# High morbidity and mortality in children with untreated congenital deficiency of leptin or its receptor

**DOI:** 10.1101/2023.03.09.23286793

**Authors:** Sadia Saeed, Roohia Khanam, Qasim M. Janjua, Jaida Manzoor, Lijiao Ning, Sharoon Hanook, Mickaël Canouil, Muhammad Ali, Hina Ayesha, Waqas I. Khan, I. Sadaf Farooqi, Giles S.H. Yeo, Stephen O’Rahilly, Amélie Bonnefond, Taeed A. Butt, Muhammad Arslan, Philippe Froguel

## Abstract

**BACKGROUND:** Biallelic pathogenic mutations in *LEP, LEPR* and *MC4R* genes controlling central leptin-melanocortin signalling cause early onset severe obesity. However, the long-term clinical outcomes of leptin signalling deficiency are unknown.

**DESIGN AND OBJECTIVES:** We carried out a retrospective cross-sectional clinical investigation of a large cohort of children with *LEP, LEPR* or *MC4R* deficiency, to evaluate the progression of the disease and its impact on morbidity and mortality.

**PARTICIPANTS:** Severely obese children from 454 consanguineous families of Pakistani origin were screened for mutations in the three genes using Sanger and exome sequencing. We identified 132 probands and 13 affected family members with homozygous pathogenic mutations in *LEP, LEPR* or *MC4R*.

**MAIN OUTCOME MEASURES:** Weight, height, and head circumference were measured by trained technicians using standardized protocols. WHO-anthro and anthroplus were used to assess BMI-standard deviation score. All affected individuals underwent detailed physical and clinical investigations by expert paediatric endocrinologist. Metabolic and oxidative stress biomarkers were measured in serum.

**RESULTS:** We report a very high mortality in children with *LEP* (26%) and *LEPR*-deficiency (9%), mainly due to recurrent pulmonary and gastro-intestinal infections. In addition, 40% of *LEP*- or *LEPR*-deficient surviving children experienced life-threatening episodes of pulmonary or gastro-intestinal infections. Oxidative stress as assessed by biomarkers, was significantly higher in *LEP* deficiency compared to the other two mutant groups.

**CONCLUSIONS:** Children with congenital deficiency of leptin or its receptor suffer a high mortality rate, and severe morbidity. Although effective therapies are available for both but as yet *(or* to-date) are not accessible in Pakistan. An appreciation of the severe impact of leptin or leptin receptor deficiency on educational attainment, morbidity and early mortality risks should spur efforts to deliver available life-saving drugs to these children as a matter of urgency.

## INTRODUCTION

Biallelic, pathogenic mutations in *LEP* encoding leptin and *LEPR* encoding its receptor, *LEPR*, that follow highly penetrant Mendelian inheritance, lead to a congenital leptin signalling deficient state, causing very early-onset severe obesity due to persistent hyperphagia. The first evidence of the pathogenic mutation in the *LEP* gene in the homozygous state resulting in leptin deficiency, was reported in two obese cousins of Pakistani origin ^1^, whereas the first inactivating homozygous mutation in *LEPR* was found in three French sisters from Algeria ^2^. Since then, 22 pathogenic mutations in *LEP* and 55 in *LEPR* have been reported, most of them in highly consanguineous populations ^3 4^.

The physiological role of leptin as a key initiator of hypothalamic neural circuitry that drives the central melanocortin pathway in the regulation of appetite is well established ^3 5^. Leptin’s binding to its receptor results in the production of α- and β-melanocyte-stimulating hormone (α-MSH and β-MSH) from pro-opiomelanocortin (POMC), peptides which act as key satiety signals by activating the melanocortin 4 receptor (MC4R) on second-order neurons. Thus, leptin-driven central melanocortin signalling is the main central regulator of energy balance, food intake and body weight.

Clinically, the initial descriptions of young patients with obesity consequent to *LEP* or *LEPR* deficiency were rather similar ^6-8^. Born with normal weight, the recessive mutation carriers gain excessive weight during infancy ^9 10^. In rodents and humans, *LEP* and *LEPR* deficiencies are associated with hyperinsulinemia, hypothyroidism, and hypogonadotropic hypogonadism resulting in delay or absence of pubertal development ^7 11 12^. Furthermore, there are some data to suggest that LEP signaling deficient individuals’ immune response may be seriously compromised with reduced number and proliferative function of T-cells and vulnerability to infections during childhood ^13-15^. However, it is noteworthy that most of the existing clinical knowledge on leptin deficiency is based on individual case reports or on small pooled data from different ethnicities ^3^. Prospective studies of genetically homogeneous individuals with leptin signaling deficiency have not been carried out so far. Thus, the long-term consequences of leptin deficiency are still unknown. Since 2012, we have constituted a unique cohort of severely obese children from consanguineous families for the study of genetic etiology of obesity in the Pakistani population (Severe Obesity in Pakistani Population [SOPP] study). We have previously reported an exceptionally high prevalence of monogenic obesity (49%) due to pathogenic mutations in known obesity associated genes, that included 33% of pathogenic homozygous point mutations or copy number variations (CNVs) in *LEP, LEPR* and *MC4R* ^16^.

Here, we present a retrospective clinical history of 92 children suffering from severe obesity due to *LEP* deficiency and 32 cases with *LEPR* deficiency, all from a single geographical region of Punjab, Pakistan. In addition, 21 cases of patients with severe obesity carrying pathogenic homozygous mutations in *MC4R*, from the same population, have been included for comparison.

## MATERIAL AND METHODS

### Study subjects

This retrospective cohort study is based on 145 cases (132 probands and 13 siblings) of monogenic obesity due to homozygous loss-of-function mutations in the *LEP, LEPR* and *MC4R* genes (Appendix p 5 Table S1). These mutations were identified through genetic screening of 454 unrelated children with severe early-onset obesity recruited to the SOPP study. The inclusion criteria were based on a body mass index standard deviation score (BMI SDS) of ≥ 3.5 and age of obesity onset 0-5 years. Where possible, other affected family members were also included in the study. In addition, a group of age-matched children negative for these mutations and of normal body weight have been included as controls.

The study was approved by the relevant institutional ethical committees, and all participants or their parents/guardians gave written informed consent. The study was conducted according to the principles outlined in the Declaration of Helsinki. A detailed interview with the patient and/or guardian, was carried out and medical history was recorded at the time of first recruitment as well as at the time of any follow-up examination. Anthropomorphic measurements were carried out and blood samples obtained (between 10 am – 12 noon), for subsequent genetic and hormonal analysis.

### Genetic analysis

Initially, all the 454 probands with severe obesity were genetically screened for mutations in *LEP* and *MC4R* genes through Sanger sequencing. DNA of the probands found negative for the mutations in these two genes, was further analysed through conventional or augmented whole exome sequencing (WES). The pathogenicity of the mutations was determined by following the American College of Medical Genetics and Genomics (ACMG) criteria. The screening methods have been described in detail elsewhere ^17 18^. Sixty-three of these affected children (56 probands and 7 siblings) were also clinically screened at a more advanced age (Appendix p 5 Table S1) but at variable intervals. Necessarily, we have included data obtained from these patients at their subsequent visit (or follow-up) as independent values in our age-related cross-sectional observations.

### Biochemical determinations

Metabolic and oxidative stress biomarkers were measured in serum. Leptin, insulin, cortisol, thyroid stimulating hormone (TSH), and 8-hydroxy-2’-deoxyguanosine (8-OHdG), were determined using commercially available ELISA kits (leptin, insulin, cortisol, TSH: Monobind, Lake Forest, USA; 8OHdG: Elabscience, Houston, USA). Serum levels of malondialdehyde (MDA) and glutathione (GSH) were determined spectrophotometrically. Samples were analysed in duplicate. The intra- and inter-assay variations were less than 11% for each assay. Random blood glucose and glycated haemoglobin A1c (HbA1c) were determined at presentation.

### Statistical analysis

Comparisons between the traits were made using Scheffe’s test. For association analysis between trait and study groups, we applied a linear regression model on different traits to assess the effect of the *LEP, LEPR* and *MC4R* mutant groups compared to the control group adjusted by gender and age. *P***-**values <0.05 were considered statistically significant. Since the number of children in the three mutant groups was not equally distributed across age (or in relation to age), the present data have also been analysed in three consecutive windows of 5 years for comparative purposes. The results were analysed with the use of the SPSS data analysis program.

The families of the three mutant groups as well as obese children with yet undiagnosed causes were approached for reporting any deaths among the affected individuals. Survival in the three groups of mutations carriers and obese controls was estimated in cases of death, and at the age of the last reported visit or contact by phone. Survival curves were estimated and plotted among the three groups of mutation carriers and obese controls, using the Kaplan-Meier method with R.

### Patient and Public Involvement

The public was not involved in the design, execution, and interpretation of this study. No participants were asked to advise on interpretation or writing of the manuscript. All the data are deidentified.

## RESULTS

### Identification of mutations in SOPP cohort

The genetic screening of 454 families from our cohort, with severe obesity, identified 132 probands (29%) and 13 family members with homozygous or compound heterozygous pathogenic variants in *LEP, LEPR* or *MC4R* (Table 1). Eighty-three children (53 males; 30 females) and nine family members (5 males; 4 females) carried 11 different homozygous pathogenic mutations in *LEP*. These included five loss of function mutations (three frameshift and two splice-site), one indel and five missense mutations. With the exception of the two missense mutations where cases presented high levels of circulating bioinactive leptin, the rest of the children presented with undetectable levels of leptin. The frame-shift c.398del.p.G133Vfs*15 was shown to be the most predominant *LEP* mutation identified in 62 patients (76%; 39 males and 23 females). This mutation was first reported in 1997 in a UK family of Pakistani origin ^1^. Interestingly, 54 probands carrying this mutation belonged to the Pakistani sub-ethnicity, *Arain*, suggesting a founder effect. For the remaining *LEP* mutations, no notable association with a particular region of origin or subethnicity was noticeable in this cohort (Table 1). Thirty-one probands (13 males; 18 females) and one family member, were identified with 21 different pathogenic homozygous or compound heterozygous variations in *LEPR*. Among these, seventeen were loss of function mutations including 5 splice-site (n = 14), four nonsense (n = 5), one start loss (n =1), three frame-shift (n = 3), one compound heterozygous (n = 1) and three CNVs with homozygous deletions of various length (44 - 52 kb). The four missense (n=4) mutations were identified in singleton case in SOPP cohort and were not identified in other 1328 children with severe obesity of the same ancestry recruited to the Genetics of Obesity Study (www.goos.org.uk), and, therefore, are likely to be private mutations. Eighteen probands (9 males; 9 females) and 3 family members (1 male; 2 females) were found to carry different homozygous mutations in *MC4R*. These included five missense, two nonsense, one frameshift and one indel variants (Table 1).

**Table 1.** Pathogenic *LEP, LEPR* and *MC4R* mutations identified in probands from SOPP cohort in relation to sub-ethnic groups.

|  | Mutation | Type of mutation | 1<sup>st</sup> reported/<br>Novel | REVEL Score | No. of probands | Sub-ethnicity |
| --- | --- | --- | --- | --- | --- | --- |
| *LEP* | c.398del (p.G133Valfs*15) | Frameshift | 1 |  | 62 | Arain (n=54), Rajput, Mughal, Gazar, Jutt, Jujklan, Tamboo, NA<sup>a</sup> (n=2) |
| *LEP* | c.313C>T (p.R105W) | Missense | 35 | 0.686 (Dam) | 1 | Sahotra |
| *LEP* | c.-28-16_-3del | Splice site | 8 |  | 2 | Mashki, Rajput |
| *LEP* | c.-29+1G>C | Splice site | 8 |  | 4 | Sheikh (n=2), Sahotra, Lotay |
| *LEP* | c.298G>A (p.D100N) | Missense | 8 | 0.610 (Dam) | 1 | Sheikh |
| *LEP* | c.309C>A (p.N103K) | Missense | 36 | 0.523 (Dam) | 2 | Arain, Rehmani |
| *LEP* | c.314G>A (p.R105Q) | Missense | 16 | 0.467 (Uncer)* | 1 | Ansari |
| *LEP* | c.350G>A (p.C117Y) | Missense | 8 | 0.33 (Uncer)* | 2 | Warraich (n=2) |
| *LEP* | c.104_106del (p.I35del) | In frame deletion | 37 |  | 5 | Rajput (n=2), Jutt (n=2), Bhatti |
| *LEP* | c.417delC (p.Y140Tfs*8) | Frameshift | 16 |  | 2 | Syed Bukhari, Jutt |
| *LEP* | c.481_482del (p.L161Gfs*10) | Frameshift | 37 |  | 1 | Jutt |
| *LEPR* | c.2396-1G>T | Splice site | 10 |  | 7 | Jutt, Rajput (n=4), Ansari (n=2) |
| *LEPR* | c.2396-2A>G | Splice site | 16 |  | 2 | Rajput, Bayi |
| *LEPR* | c.704-1G>A | Splice site | 16 |  | 3 | Ansari, Bhatti, Khansani |
| *LEPR* | c.2213-3C>G | Splice site | 16 |  | 1 | Kakazai |
| *LEPR* | c.994+2T>C | Splice site |  |  | 1 | Mughal |
| *LEPR* | c.2T>C (p.?) | Start loss | 16 |  | 1 | Awan |
| *LEPR* | c.1674G>A (p.W558*) | Nonsense | 10 |  | 2 | Siddiqui, Sheikh |
| *LEPR* | c.133_136dup p.(Tyr46*) | Nonsense | Novel |  | 1 | Lalikher |
| *LEPR* | c.2114G>A (p.W705*) | Nonsense | 16 |  | 1 | Dogar |
| *LEPR* | c.489T>A (p.Y163*) | Nonsense | Novel |  | 1 | Barfizai |
| *LEPR* | c.1810T>A (p.C604S) | Missense | 16 | 0.609 (Dam) | 1 | Gujar |
| *LEPR* | c.2627C>T (p.P876L) | Missense | 16 | 0.647 (Dam) | 1 | Mughal |
| *LEPR* | c.40G>A (p.E14K) | Missense | 16 | 0.116 (Ben)<sup>‡</sup> | 1 | Rehmani |
| *LEPR* | c.2153A>G (p.N718S) | Missense | 16 | 0.665 (Dam) | 1 | Jutt |
| *LEPR* | c.2899_2900insAT(p.A967Dfs*7) | Frameshift | 16 |  | 1 | Jutt |
| *LEPR* | c.3268_3269del (p.S1090Wfs*6) | Frameshift | 38 |  | 1 | Arain |
| *LEPR* | c.1738delG(p.E580Kfs*37) | Frameshift | 16 |  | 1 | Arain |
| *LEPR* | c.2875T>A (p.R959*) & c.2872G>A (p.E958K) | Compound heterozygous | 16 |  | 1 | Gujar |
| *LEPR* | ~44 kb homozygous deletion including exon 3 to 20 of *LEPR* | CNV | 16 |  | 1 | Gujjar |
| *LEPR* | ~61 kb homozygous deletion including exon 3 to 6 of *LEPR* | CNV | 8 |  | 1 | Rajput |
| *LEPR* | 52 kb homozygous deletion | CNV | 16 |  | 1 | Rajput |
| *MC4R* | c.633_636delCTAT (p.Y212Sfs*5) | Frameshift | 16 |  | 1 | Khokhar |
| *MC4R* | c.47G>A (p.W16*) | Nonsense | 39 |  | 3 | Jutt, Rajput (n=2) |
| *MC4R* | c.63_64del (p.Y21*) | Nonsense | 16 |  | 2 | Arain (n=2) |
| *MC4R* | c.601_612 del<br>TTCTTCACCATG<br>(p.F201_M204del) | In frame deletion | 16 |  | 1 | Rehmani |
| *MC4R* | c.206T>C (p.I69T) | Missense | 40 | 0.592 (Dam) | 1 | Jutt |
| *MC4R* | c.482 T>C (p.M161T) | Missense | 40 | 0.488 (Uncer) | 3 | Pahan (n=2), Memon |
| *MC4R* | c.493C>T (p.R165W) | Missense | 41 | 0.501 (Dam) | 4 | Arain (n=2), Jutt<br>Sayal, Lashari |
| *MC4R* | c.811T>C (p.C271R) | Missense | 30 | 0.844 (Dam) | 1 | Bhatti |
| *MC4R* | c.947 T>C (p.I316S) | Missense | 42 | 0.442 (Uncer) | 2 | Rajput, Sheikh |

<sup>a</sup>Not available, Dam: damaging, Uncer: Uncertain, Ben: benign
LEP: NM\_000230.3; LEPR:002303.6; MC4R:005912.3
<sup>†</sup> Located at essential splice-site
\*Undetectable levels of leptin hormone
LEP: NM\_000230.3; LEPR:NM\_002303.6; MC4R:NM\_005912.3

### Clinical Phenotypes

The most prominent and ubiquitous characteristic of children identified with biallelic pathogenic mutations in *LEP, LEPR* and *MC4R* genes was the onset of severe obesity and hyperphagia at a very young age–they lacked satiety and were constantly in demand of food. If denied, they craved for more food by crying and turning aggressive. the majority of *LEP* and *LEPR* deficient children displayed excessive weight gain and hyperphagia that was noticed at a far earlier age in postnatal life (before 1 year of age; mean age in *LEP* deficient: 0.3±0.04 year [n = 83] and in *LEPR* deficient (0.9 ± 0.1 year [n = 27]), as compared to those in children with MC4R deficiency (mean age: 4.0±3.3 years [n = 20]).

Children deficient for *LEP* or *LEPR*, presented a more precocious and profound hyperphagia than those of children deficient for *MC4R* and with frequent aggressive demand for food during the day and whilst waking up at night. The craving for food was reported relatively less intense and frequent in children with *MC4R* deficiency. At the time of clinical examination, 84% of *LEP*-deficient, 79% *LEPR*-deficient children, and 61% of *MC4R*-deficient children suffered from severe hyperphagia, whereas in the remaining affected subjects, the craving for food though frequent was reported as less severe or moderate (Figure 1). It is noteworthy that with advancing age no exacerbation of severity of hyperphagia was observed. On the contrary, a considerable variation in demand for food with time, was observed in *LEP*- or *LEPR*-deficient children. This may partly be due to parents’ care and efforts to divert the attention of the child’s reward system to other activities of choice.

**Figure 1.**
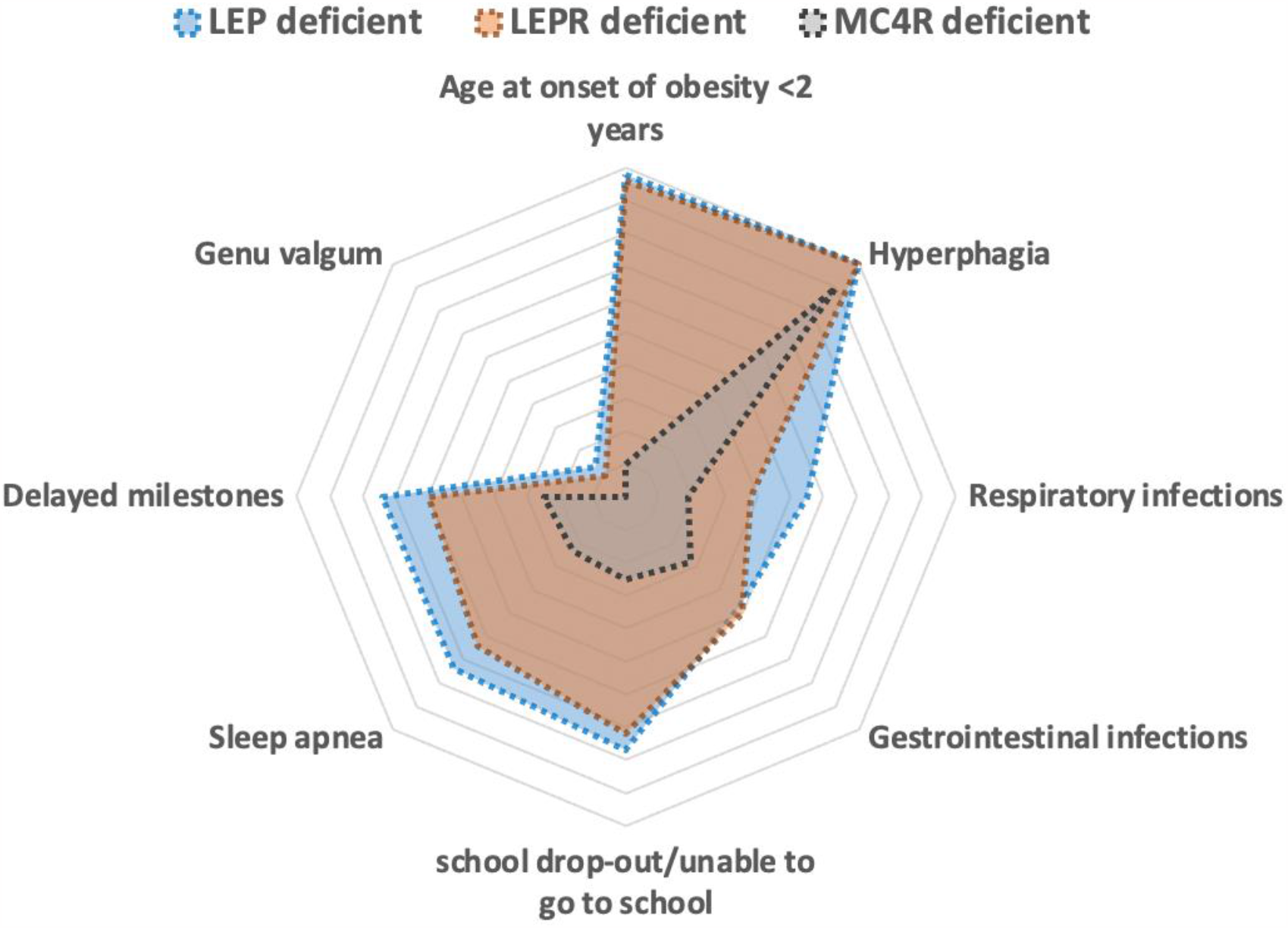
Radar chart representing the severity of the obesity-associated risks in children with *LEP* and *LEPR* deficiency compared to those with *MC4R* deficiency.

No dysmorphic features or developmental deformities were observed in any of the obese probands. However, acanthosis nigricans was often present in the three mutation carrier groups and was most predominant as dark velvety patches of skin around the neck (Appendix p 16 Figure S1a). Figure 1a). Furthermore, prominent skin folds braced by the subcutaneous fat were invariably present on the forearms and trunk region of children deficient for *LEP* or *LEPR* at an early stage of life but were less conspicuous in *MC4R*-deficient age-matched individuals (Appendix p 16 Figure S1b).

#### Morbidity

The majority of the *LEP*- or *LEPR*-deficient children (55% and 38%, respectively) suffered from recurrent episodes of serious respiratory afflictions such as pneumonia and upper respiratory tract infections. Also, the majority of leptin and leptin receptor deficient children (74% and 64%, respectively) were assessed as hypoxic. The second most common complication observed is related to gastro-intestinal infections often leading to severe diarrhoea. About 40% of the living children with *LEP* or *LEPR* deficiency had one or more hospital admissions for intensive care management of these complications. In addition, genu valgum was identified in 13% of children deficient for *LEP* and 9% of children with LEPR deficiency but was not reported in the MC4R deficient group (Figure 1). Vision refractive errors were reported in 16% of probands with *LEP* deficiency, but not in the other two groups.

#### Mortality

The deaths were reported in 26% of *LEP*-deficient children and 9% of children with *LEPR* deficiency (Figure 1, (Appendix p 6 Table S2). In the majority of these cases, the cause of death was diagnosed as respiratory failure due to pneumonia or other respiratory infections and the second most common cause of death was severe diarrheal episodes. No deaths have so far been reported in *MC4R*-deficient children from this cohort. The Kaplan Meier survival estimates demonstrate a remarkably low survival rate in children with leptin signaling deficiency (*P* < 0.05; Figure 2).

**Figure 2.**
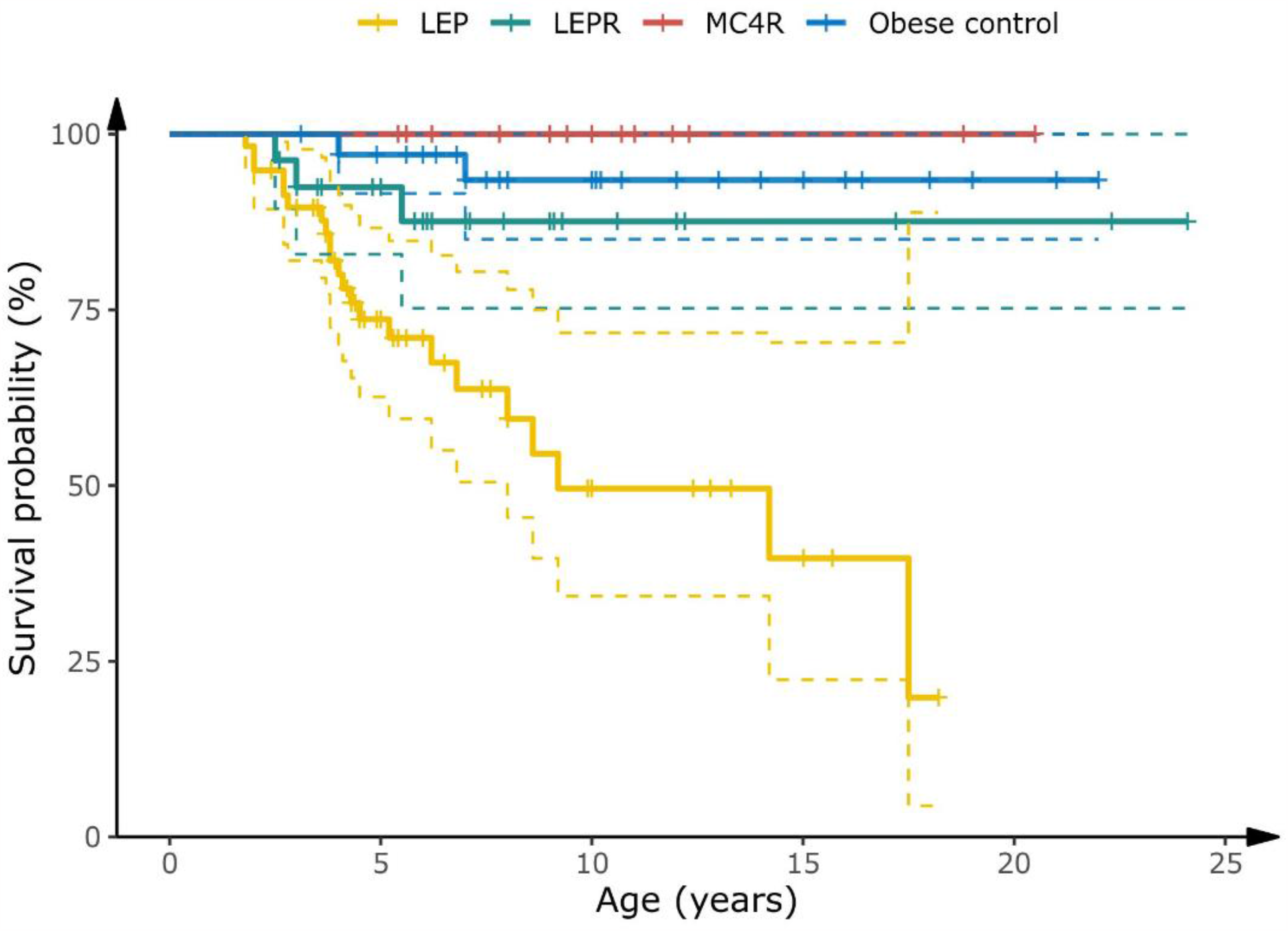
Survival curve of children with *LEP, LEPR* and *MC4R* deficiencies and severely obese children negative for mutations in known obesity genes (obese controls) (Kaplan-Meier Group Log-rank *p* < 0.05).

#### Physical activity and social behaviour

The gross motor development milestones were noticeably delayed in 74% of children with *LEP*, 60% *LEPR* and 25% *MC4R* deficiencies, presumably and partly due to excessive adiposity. Thus, the ability to independently sit and walk was acquired by these children at a much later stage, compared to their non-obese siblings or to age-matched normal subjects. Aggressive behaviour in majority of children with *LEP* and *LEPR*-deficiency (84% and 50% respectively) was reported towards their siblings and peers, but not towards their parents. The relative inability to learn and remember new things was variable in affected children but was more evident in children with *LEP* and *LEPR* deficiency (77% and 72% respectively) as compared to those of *MC4R* deficient children (25%). Overall, only a small percentage of the *LEP*-deficient (22%) and *LEPR*-deficient (27%) children over the age of 5 years were reported to be attending school compared to 75% of *MC4R*-deficient children. Furthermore, those children who initially attended school, very often dropped out prematurely mainly due to their inability to keep in-step with their peers in studies, lack of integration and acceptance by their peers, frequent health problems, and social pressures (Figure 1). A lag in learning abilities in children with *LEP* and *LEPR* mutations probably indicates an age-related delay in cognitive ability. However, no attempt was made to quantitatively assess cognitive limitations in these patients.

#### Body growth

Longitudinal growth data in relation to age in the three analyzed groups, and in control subjects with normal body weight, from age 1-15 years are presented in Appendix p 17 Figure S2 and Appendix p 7 Table S3. The mean height was significantly greater (*P* < 0.05) in prepubertal children with *MC4R* deficiency than in the other two study groups whereas no significant differences in growth were found between *LEP* and *LEPR* deficient subjects. The estimated linear growth in *MC4R* deficient children was also significantly increased compared to the age and gender-matched controls (8.6±2.2; *P* = 0.0001). We observed a similar trend (but statistically non-significant) with *LEP* and *LEPR* deficient subjects (1.7±1.6 and 2.2±1.8 respectively) (Appendix p 7 Table S3). No remarkable differences were found in longitudinal growth in individuals carrying mutations and the control subjects, during the first 10 years of life (Appendix p 7 Table S3). However, we noticed a trend towards an increased growth in individuals with *MC4R* deficiency in children ≥10-years old, compared to age-matched *LEP* and *LEPR* deficient and also to the control subjects. Age related body mass data are summarized in Appendix p 18 Figure S3, Appendix p 7 Tables S3 and Appendix p 10 S6.

### Metabolic Characteristics

The overall metabolic characteristics of children with mutations in the three genes and those with normal body weight, are presented in Table 2. As expected, insulin levels were significantly higher in the three mutation carrier groups compared to control values and increased progressively with advancing age (Appendix p 11 Table S7). At the age of 10-15 years, hyperinsulinemia appeared milder in the *LEP* deficient group as compared with the LEPR and MC4R deficient patients (mean 49 μIU/ml *versus* 104 μIU/ml and 66 μIU/ml in *LEP, LEPR*, and *MC4R* deficient children, respectively (range of insulin assay (0-300 μIU/ml).

**Table 2.** Endocrine and glycaemic characteristics of children with *LEP, LEPR* and *MC4R* mutations and normal controls.

| Characteristic | *LEP* deficient | *LEPR* deficient | *MC4R* deficient | Controls |
| --- | --- | --- | --- | --- |
| Leptin (ng/ml) | 0.1 ± 0.0<sup>b, c</sup> | 31.9 ± 2.7<sup>a, d</sup> | 26.7 ± 3.5<sup>a, d</sup> | 3.1 ± 0.1<sup>b, c</sup> |
| (N) | (108) | (49) | (26) | (68) |
| Insulin (uIU/ml) | 24.6 ± 2.3<sup>b, c, d</sup> | 41.8 ± 5.9<sup>a, d</sup> | 49.0 ± 10.3<sup>a, d</sup> | 6.9 ± 0.5<sup>a, b, c</sup> |
| (N) | (111) | (49) | (26) | (61) |
| Cortisol (ug/dl) | 16.2 ± 0.8<sup>b, c, d</sup> | 13.3 ± 0.7<sup>a, d</sup> | 10.9 ± 0.8<sup>a</sup> | 9.4 ± 0.3<sup>a, b</sup> |
| (N) | (111) | (49) | (26) | (61) |
| TSH (uIU/ml) | 2.2 ± 0.1 | 2.3 ± 0.2 | 2.1 ± 0.2 | 2.2 ± 0.1 |
| (N) | (112) | (49) | (26) | (61) |
| HbA1C | 5.2 ± 0.1<sup>d</sup> | 5.0 ± 0.1<sup>c, d</sup> | 5.9 ± 0.5<sup>b, d</sup> | 4.2 ± 0.1<sup>a, b, c</sup> |
| (N) | (29) | (19) | (7) | (40) |
| RBG (mg/dl) | 95.7 ± 3.2 | 99.7 ± 3.7 | 109.1 ± 7.8 | 102.8 ± 2.6 |
| (N) | (30) | (23) | (15) | (40) |
| T4 (µg/dl) | 8.4±0.6 | 7.0±0.5 | 7.6±0.9 | 8.1±0.3 |
| (N) | (23) | (11) | (15) | (27) |
| T3 (ng/ml) | 1.8±0.1 | 1.9±0.2 | 1.6±0.1 | 1.5±0.1 |
| (N) | (23) | (11) | (15) | (27) |

Superscript letters indicate statistically significant P < 0.05 (Scheffe's multiple test); <sup>a</sup> vs *LEP* deficient, <sup>b</sup> vs *LEPR* deficient, <sup>c</sup> vs *MC4R* deficient, <sup>d</sup> vs controls
Data represent Mean ± SEM

Leptin concentrations in *LEPR* deficiency tended to be higher in the first 5 years of life than those of the *MC4R* deficient children (*P* < 0.05) (Appendix p 12 Table S 8). As anticipated, serum leptin concentrations were below the sensitivity level of the assay in *LEP*-deficient children with the exception of two individuals, with the p.N103K mutation and one individual with p.D100N mutation, in *LEP* (0.1-5.0 years of age) These cases present high levels of circulation of immunoreactive, but bioinactive leptin protein (mean: 49 ng/ml) as also reported previously ^19^.

Mean cortisol levels (am) were significantly higher (*P* < 0.05) in *LEP* deficiency than those in the other two groups but tended to decline with age (Appendix p 13 Table S9). The mean cortisol levels in *MC4R* deficiency were indistinguishable from the controls. Remarkably, mean serum TSH, T4 and T3 concentrations were within the normal range in *LEP, LEPR* and *MC4R* deficient children, and were not different from those of normal subjects (Table 2, Appendix p 14 Table S10). Mean HbA1c ranged between 5.0-5.9 in the three mutant groups compared to a mean of 4.2 in the control group (Table 2). Mean random blood glucose levels (RBG) were within the normal range in the three groups of mutation carriers and comparable to control values (Table 2), except for one individual with MC4R deficiency (RBG = 199 mg/dl).

### Oxidative stress

Peripheral levels of malondialdehyde (MDA) and 8-OHdG, critical biomarkers of oxidative stress that occurs as a consequence of an imbalance between the formation of free oxygen radicals and inactivation of these species by antioxidant defence system, and of glutathione (GSH), an antioxidant biomarker, were measured in a restricted number of probands (Table 3) Serum levels of MDA and 8-OHdG were significantly higher and of GSH lower in *LEP* and *LEPR* deficient individuals compared to those with *MC4R* deficiency (*P* < 0.05). Circulating levels of MDA and 8-OHdG were maximal in *LEP* deficient children.

**Table 3.**
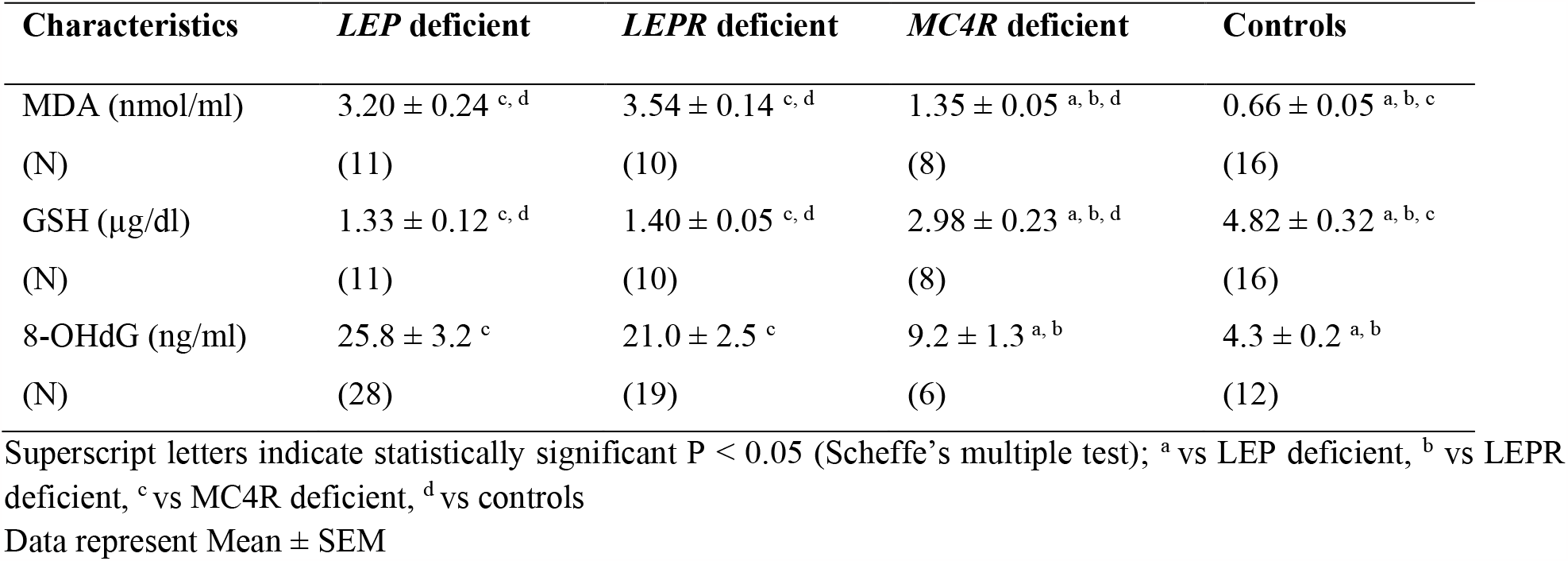
Serum levels of stress biomarkers in children with *LEP, LEPR* and *MC4R* mutations.

## DISCUSSION

Our study demonstrates the dramatic mortality rate in obese children with LEP deficiency and to a lesser extent in children with LEPR deficiency. While increased susceptibility to infection and mortality have been reported in case reports of these conditions previously ^7 20^, this large clinical series of children definitively establishes the severe clinical impact of these conditions. In contrast, no severely obese child with MC4R deficiency of same ethnic origin died during the study. Indeed, all children with severe obesity in this study belong to the same geographical region and share similar socioeconomic status (low middle-economy). Previous studies indicate that the country’s disease burden may be overtly influenced by the socio-economic index^24.^ In LEP signaling deficiency, the mortality rate may even be underestimated, as ∼30% of the families could not be contacted following the initial presentation. Health records indicated that deaths of the children were mostly due to respiratory and gastrointestinal infections. These effects may be due to lowered immunity and possibly a lack of a permissive effect of leptin on immune cells ^13 21^.

Previously, clinical investigation of three leptin deficient patients demonstrated an increased frequency of infections predominantly of the respiratory tract when comparing with their wild-type siblings. T-cell number and function were impaired in the subjects with a reduced number of CD4+ T cells, reduced T-cell proliferation and Th2 cytokine response but no gross abnormalities related to thermoregulation were reported ^13^. These findings of T-cell responsiveness were consistent with observations in ob/ob mice ^22^. Therapeutic doses of recombinant leptin normalized the T cell function^13^. These findings suggest that leptin is a crucial metabolic signal that allows activation and proliferation of T-cells thus linking nutritional status, cellular metabolism, and immunity ^23^.

Another outcome of this investigation is the observation that as many as 40% of living young patients with LEP/LEPR deficiency had experienced serious health problems posing a life-threatening risk and necessitating hospitalization for intensive care management. According to a recent UNICEF (The United Nations Children’s Fund) report, the mortality in children under five years of age (excluding neonatal mortality which constitute more than half of the under five deaths in the country) is estimated at 2.5% in Pakistan, for the year 2020 (ref: Monitoring the situation of children and women. Pakistan, Key demographic indicators 2020. (https://data.unicef.org/country/pak/). Our data unequivocally demonstrate that leptin deficiency leads to a remarkably higher mortality rate. presumably due to lowered immunity leading to high risk of infections in these children. Also, according to the Global Burden of Diseases, Injuries, and Risk Factors Study (GBD) 2019, neonatal disorders, congenital defects, diarrhoeal diseases and respiratory infections, have been shown to be among the top ten main causes of premature mortality and years of life lost, in Pakistan ^24^.

Although severe obesity has been considered as a long-term risk factor of early morbidity, disability, and premature death in adulthood, this is the first time, to our knowledge, that a well-defined non-syndromic form of genetic obesity in childhood has been unambiguously shown to be so dramatically life threatening. Indeed, more common forms of severe early onset obesity are associated with metabolic and cardio-vascular disease as well as with some cancers later in life, but not with high infection rates (with the recent exception of COVID19 in relatively young adults. Earlier research on leptin signalling deficiency in human and animals suggested impaired immunity that may contribute to the severity of common respiratory and digestive infections ^13^. Although we were not able to carry out direct evaluation of humoral and cellular immunity in our obese children, we found a high systemic oxidative stress level and a marked depletion of the antioxidant, GSH, in obese carriers with homozygous loss-of-function mutations in *LEP* or *LEPR* genes. In comparison, the oxidative stress level was found to be lower in patients with MC4R deficiency. Earlier studies suggested a positive correlation between body mass index and levels of oxidative stress^24 25^ but here we demonstrate that the level of oxidative stress related to obesity may differ even between well-defined conditions of genetic obesity. In spite of a close phenotypic identity between *LEP* and *LEPR* mutation carriers, morbidity in LEP deficient children is surprisingly much higher than that of LEPR deficient patients. We also found a higher index of oxidative stress and DNA damage in LEP deficient children compared to those with LEPR deficiency, suggesting a more severe impairment in cellular or humoral immunity of unknown nature.

The very early onset of weight gain due to severe hyperphagia in LEP and LEPR deficient subjects, is well documented ^6^, and in this respect we confirm that LEP and LEPR deficient children are indistinguishable ^7 16 21^. In addition, our ethnically homogeneous study unambiguously shows that the onset of severe obesity is ∼3 years earlier in both LEP and LEPR deficiencies compared to MC4R deficient children.

Here we also document the high incidence of delay in learning possibly due to intellectual impairment, and aggressive behaviour in LEP and LEPR deficiencies, that appear to be milder or absent in MC4R deficient children. This is also evidenced by the observation that only ∼25% of LEP or LEPR deficient children could attend school after the age of 5 years compared to 75% of children with MC4R deficiency. Also, long-term schooling was nearly impossible for LEP/LEPR deficient children. This other dramatic outcome is further supported by our recent study on a group of severely obese teenagers from the same geographical region and with the same level of consanguinity (mean BMI = 37 kg/m^2^; mean Age = 18 years) who were regularly attending school. Remarkably, this cohort did not include a single subject carrying a mutation in *LEP* gene ^17^.

LEP deficient *ob/ob* and LEPR deficient *db/db* mice have decreased linear body growth ^26^. In human, previous reports on linear growth due to defective LEP or LEPR signalling have been controversial ^2^, with reports of both accelerated ^27 28^ and diminished^2 29^ linear growth. Here, our data in a much larger cohort demonstrate no remarkable differences in linear growth in LEP or LEPR deficient children compared to age-matched controls up to the age of 15 years. In the present investigation, we observed a significantly increased mean linear growth in MC4R deficient children as compared to the other two groups of mutation carriers. Previously, MC4R deficiency has also been associated with increased linear growth due to enhanced osteogenesis ^30^.

Hyperinsulinemia, though significantly more pronounced in LEPR and MC4R deficient subjects compared to those with LEP deficiency, increased with age in all three mutant groups. Raised HbA1c levels (but still within the normal range) were found in MC4R deficient children, but to a lesser extent in LEP and LEPR deficient patients. Consequently, none of these children had thus far developed a risk of diabetes. The euglycemic condition in affected children in this study is in concordance with previous observations ^30^.

Hypercortisolemia was more prominent in children with LEP deficiency compared to those with LEPR deficient patients whereas cortisol levels were in the normal range in MC4R deficiency and indistinguishable from those of the controls, as previously reported ^8^. This corresponds to the high cortisol levels seen in *ob*/*ob* and *db*/*db* mice ^31 32^. Observations on thyroid function in subjects with these mutations were again conflicting ^13 33^. Notably, in this large study, serum TSH, T4 and T3 levels were within the normal range in all the three mutation carrier groups thus excluding thyroid abnormalities in children in this study. However, here no attempt was made to estimate free T4 levels.

### Strengths and limitations of the study

The main strength of the study is that it is based on an exceptionally large number of individuals with leptin signaling deficiency - a globally rare event, from a single geographical region, This has enabled us to clinically follow and assess the age-related progression of the disease, morbidity and mortality, during childhood to adolescent stage.

This investigation has certain limitations. Firstly, it is necessarily a cross sectional study and lacks the advantage of an organized follow-up regimen. A regular follow-up was not possible because of logistic problems as a sizeable proportion of affected families especially those residing in rural and remote areas of the province/country did not respond to a follow-up call or could not be contacted second time.

## CONCLUSIONS

In summary, comparative data from this retrospective cross-sectional study indicate a distinctly higher level of morbidity in LEP and LEPR deficient obese children compared to those with homozygous loss-of-function mutations in the *MC4R* gene. Hormone replacement therapy with the leptin analogue – Myalept, for treatment of leptin deficient subjects and MC4R agonist – setmelanotide for treatment of LEPR deficient subjects ^34^ though available in many developed countries, are unfortunately lacking in Pakistan - a country with the world’s highest recorded prevalence of LEP signalling deficiency. The fact that a sizeable population of children is failing to achieve normal educational development, becoming seriously ill and dying prematurely as the result of a deficiency in hormonal signalling for which relatively simple peptide treatments are readily available, highlights serious flaws in the global system through which drugs are developed and made available to those who most need them.

## Data Availability

All data produced in the present work are contained in the manuscript.

## CONTRIBUTORS

SS, SOR, MA and PF conceptualized the study. SS, MA and PF wrote the first draft of the manuscript. JM, ISF, GSHY, SOR and AB contributed in reviewing and editing the first draft. SS, RK, QMJ. and MA collected samples and performed biochemical analysis. S.S, A.B, MA and PF performed analysis of the genetic data. SS, RK, QMJ, LN, SH, MC, AB and MA performed statistical analysis. JM, MAli, HA, WIK and TAB identified and recruited obese families. SS, MA and PF are the guarantors of this work and, as such, had full access to all the data in the study and take responsibility for the integrity of the data and the accuracy of the data analysis. All authors contributed to the revision process of the manuscript and approved the final version of the manuscript.

## DECLARATION OF INTERTERST

No potential conflicts of interest relevant to this article were reported.

## DATA SHARING

Qualified researchers can request patient-level data, including the study protocol with any amendments and blank patient information form. Patient-level data will be anonymised, and study documents will be redacted to protect the privacy of the trial participants.

## ETHICAL APPROVAL

This study involves human participants and was approved by the Children’s Hospital and Institute of Child Health (now Children’s Hospital and UCHS) Ethical approval letter no. 5178/PH. Participants gave informed consent to participate in the study before taking part.

## ACKNOWLEGDMENTS

We thank Frédéric Allegaert, Timothée Beke and Stefan Gaget for technical assistance. This work was supported by funding from the Medical Research Council (MRC) MR/S026193/1 (P.F.) and the Pakistan Academy of Sciences (M.A). S.S is supported by a project grant from the Medical Research Council (MR/S026193/1). Further support was provided by the National Center for Precision Diabetic Medicine – PreciDIAB, which is jointly supported by the French National Agency for Research (ANR-18-IBHU-0001), by the European Union (FEDER), by the Hauts-de-France Regional Council and by the European Metropolis of Lille (MEL). We thank the patients and families for their participation in the study.

